# Rationale, protocol and baseline characteristics of the metabolome, microbiome, and dietary salt intervention study (MetaSalt)

**DOI:** 10.1101/2021.03.06.21252976

**Authors:** Zengliang Ruan, Jianxin Li, Fangchao Liu, Jie Cao, Shufeng Chen, Jichun Chen, Keyong Huang, Yaqin Wang, Hongfan Li, Yan Wang, Xue Zhongyu, Laiyuan Wang, Jianfeng Huang, Dongfeng Gu, Xiangfeng Lu

## Abstract

High sodium intake has been recognized as an important risk factor for hypertension, but the role of gut microbiota composition and metabolomic profiles in the association between dietary sodium intake and blood pressure (BP) is uncertain. The metabolome, microbiome, and dietary salt intervention study (MetaSalt) study was conducted to investigate whether low- and high-dietary sodium intake influences BP by changing the microbial and metabolomic profiles. This is a family-based, multicentre intervention study conducted in four rural field-centres across three provinces in rural Northern China. Probands with untreated prehypertension or stage-1 hypertension were identified through a community-based BP screening, and their family members included siblings, offspring, spouses and parents were subsequently included. During the dietary intervention, low-salt and high-salt diets were provided free of charge to all participants. A total of 529 participants in four field centres were included in our study, with a mean age of 48.1 years old, and about 36.7% of them were male, 76.5% had a middle school (69.5%) or higher (7.0%) diploma, 23.4% had a history of smoke, 24.4% were current drinkers. The mean systolic and diastolic BP levels in the baseline were 129.54 mm Hg and 81.02 mm Hg for all participants, and significantly decreased during the low-salt intervention and increased during the high-salt intervention. Our study is well placed to check the impacts of dietary sodium intake on microbial and metabolomic profiles, which will have important implications for discovering the mechanisms in the development of hypertension and subsequent cardiovascular disease.

## Introduction

Hypertension, as a major risk factor for cardiovascular disease and the leading cause of death and disability, accounted for 19.2% of all deaths and 9.3% of all disability-adjusted life years (DALYs) worldwide in 2019 [1-3]. The positive and significant relationship between dietary salt (sodium) intake and blood pressure (BP) has been demonstrated by a series of prospective studies, which suggested that dietary salt intervention could be an effective and low-cost measure to control hypertension [4, 5]. However, BP changes in response to sodium intakes vary among different populations, which is a physiological trait called salt sensitivity, and the underlying mechanisms remain incompletely understood [6].

Previously, we initiated the Genetic Epidemiology Network of Salt Sensitivity (GenSalt) study, which has identified several susceptibility genes that might play important roles in determining individual BP responses to dietary sodium intake, and provided substantial evidence to support a genetic basis for the variation in the BP response to salt [6, 7]. For example, findings from the GenSalt study have suggested that genetic variants in the endothelial system, sympathetic nervous system, and ion and water channels, could influence the sodium sensitivity of BP [8-12]. Moreover, recent advances have revealed that dietary sodium intake is associated with changes in gut microbiota and circulating or urinary metabolites, and these changes might further influence BP through different pathways [13, 14]. For example, prior research findings have suggested that excess dietary salt was associated with increases in several intestinal microbes that caused microbiome-induced inflammation, which would further result in higher BP in humans and mice [15]. Several metabolites that could be affected by the interaction of intestinal microbiota and dietary sodium intakes, such as triglycerides, linoleate, lipopolysaccharide, short-chain fatty acids (SCFAs), dihomolinolenate, bile acids, trimethylamine-N-oxide (TMAO), sphingomyelins and uremic toxins, were identified to be involved in BP regulation, cardiovascular health and disease [16-19]. Fumarase, an enzyme in the tricarboxylic acid cycle, was also found to be significantly reduced in the Dahl salt-sensitive (SS) rat, a widely-used animal model of human salt-sensitive hypertension, which suggested that abnormalities in cellular intermediary metabolism might affect the salt-sensitivity of BP [20]. To our knowledge, previous findings on the associations of dietary sodium intake with the microbial and metabolomic profiling were mainly concluded from animal experiments or observational studies [16, 21-23], but only a few intervention trials were performed to verify their causal relationship in human, and none of these trials was conducted in the Chinese population [24, 25].

Accordingly, the metabolome, microbiome, and dietary salt intervention study (MetaSalt) study was designed to identify alternations in the microenvironment (i.e., oral and gut microbiota and their metabolites) and plasma metabolites induced by low and high dietary salt intakes.

## Methods

### Study design

The MetaSalt study is a family-based, multicentre dietary salt intervention trial, conducted in rural areas of Northern China. The study was registered in the Chinese Clinical Trial Registry (ChiCTR) database on August 15, 2019 (registration number: ChiCTR1900025171). The protocol was developed following the Declaration of Helsinki and approved by the Ethics Committee of Fuwai Hospital [26], and written informed consent was obtained from all participants.

### Study site and population

The MetaSalt study was carried out in four rural field-centres in Hebei (Xinle city and Nangong city), Shanxi (Yu County) and Henan (Changge County) provinces in rural Northern China (Figure 1). These study sites were selected in terms of their similar features in the geographical, environmental, cultural, and habitual diet of the population. The study participants were recruited from families with high risk for the development of hypertension because they are like to be sensitive to dietary sodium intake [6, 27].

**Figure 1.**
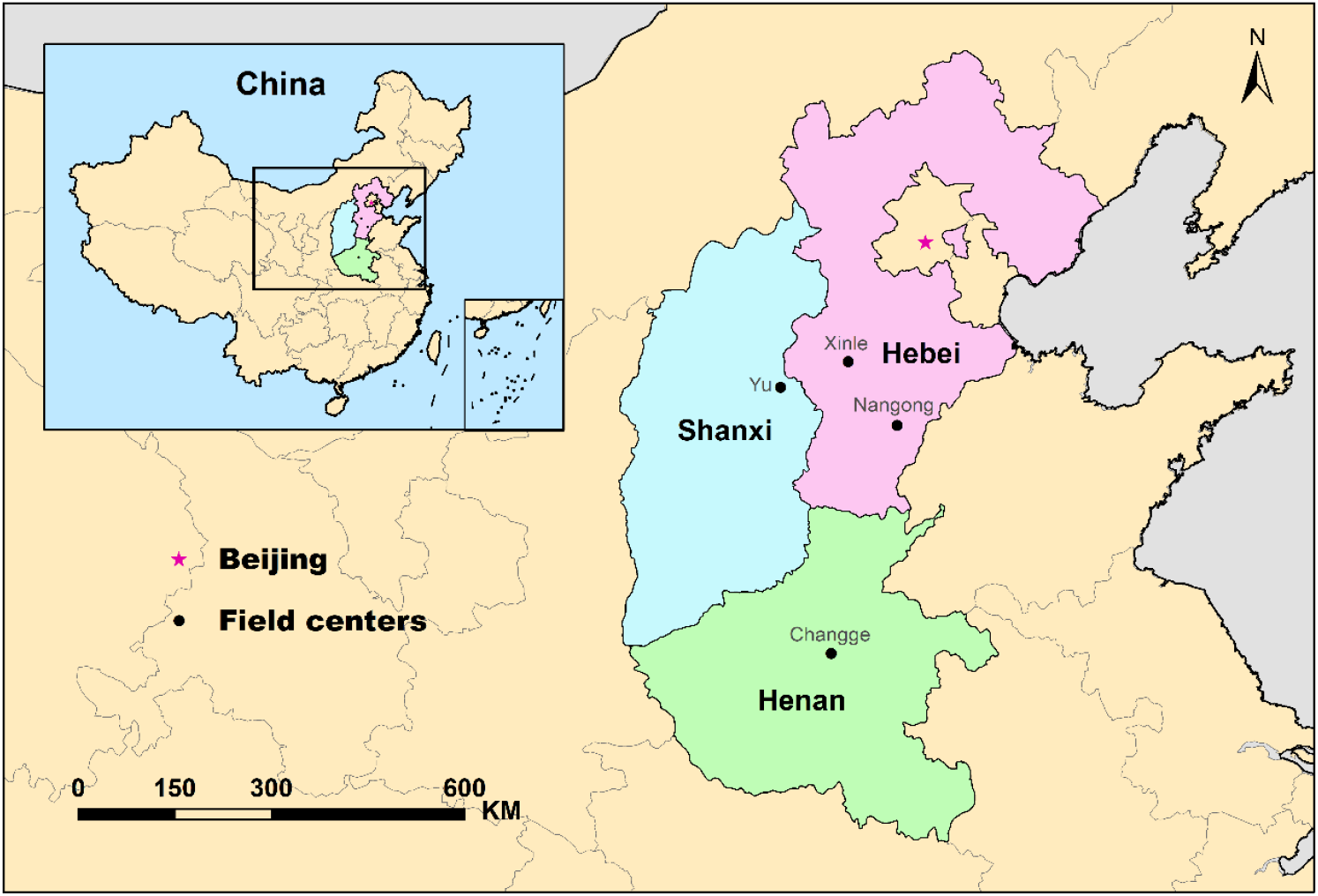
Geographical location of the study sites.

### Participants

The recruitment of the MetaSalt study was conducted from August to November of 2019. All participants of the MetaSalt study were rural residents aged 18–60 years from the four study sites, and they are of the ethnic majority group in China (i.e., Han nationality). According to the inclusion and exclusion criteria (Table 1 and 2), the potential probands were first identified by both a short questionnaire and community-based BP screen in the selected villages. Furthermore, their family members (parents, spouses, siblings and offspring) were selected based on the criteria (Table 1 and 2). To confirm the eligibility of each participant, we carried out the second round of BP screens on the next day among the identified subjects. Finally, the confirmed participants were invited to sign an informed consent before they were officially enrolled.

**Table 1.** Inclusion criteria for the participants of the MetaSalt study.

|  |
| --- |
| ● Aged 18 to 60 years; |
| ● Without secondary hypertension; |
| ● Blood pressure: |
| ◆ Probands: Systolic BP 130 to 159 mmHg and/or diastolic BP 85 to 99 mmHg (mean value of three measurements); |
| ◆ Family members: Systolic BP <160 mmHg and diastolic BP <100 mmHg (mean value of three measurements); |
| ● No antihypertensive drugs or drugs affecting BP were taken within 1 month before the screening visit; |
| ● Negative in the urine protein test and normal blood glucose level (fasting blood glucose < 7.0 mmol/L and postprandial blood glucose < 11.1 mmol/L); |
| ● Family members: living in the same or neighbouring village with the proband; |
| ● Provided informed consent. |

**Table 2.** Exclusion criteria for the participants of the MetaSalt study.

|  |
| --- |
| ● Use antibiotics or antihypertensive drugs within 1 month before the screening visit;<br>● Defecation interval time $\geq 3$d;<br>● Stage-2 or more serious hypertension patients (systolic BP $\geq 160$ mm Hg and/or diastolic BP $\geq 100$ mm Hg);<br>● History of cardiovascular disease, including coronary heart disease, heart failure, stroke, peripheral arterial disease, etc.;<br>● Having kidney disease (positive in the urine protein test);<br>● Having diabetes (fasting blood glucose $\geq 7.0$ mmol/L and postprandial blood glucose $\geq 11.1$ mmol/L) or use of insulin or oral hypoglycaemic drugs;<br>● History of peptic ulcer disease or liver disease which required treatment during the past 2 years;<br>● Currently pregnant;<br>● Drink alcohol almost every day;<br>● Currently adhering to a low-sodium diet;<br>● Unwilling to sign the informed consent. |

### Intervention

The MetaSalt study was conducted for 25 consecutive days, including two days of screening, three days for baseline observation, ten days for low dietary sodium intervention and another ten days for high dietary sodium intervention among the participants. The flowchart of the intervention strategy was presented in Figure 2. During the baseline observation, all participants kept their usual diet, and the intervention was started on the sixth day, all participants received a low-salt diet (3 g/d of salt or 51.3 mmol/d of sodium) for 10 days (days 6 to 15), followed by a high-salt diet (18 g/d of salt or 307.8 mmol/d of sodium) from days 16 to 25. During the intervention period, all participants took 3 meals per day (i.e., breakfast, lunch and dinner) at specified canteens, which were cooked by the study staff without salt. The sodium intake of each participant was strictly controlled through pre-packaged salt that was added to the foods by study staff. In addition, the daily recipes were determined by professional dietitians, and although dietary salt intake was equal for all subjects, their dietary total energy intake was in accordance with the baseline information which was estimated by a food frequency questionnaire.

**Figure 2.**
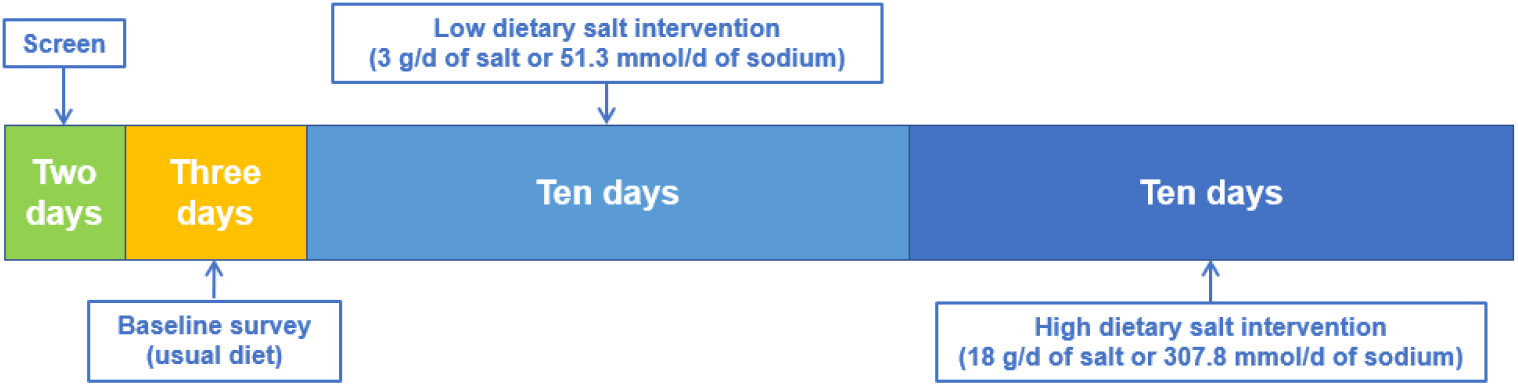
The flowchart of this study.

### Data collection and measurements

Data collection, measurements, and sample collection at every stage are summarized in Table 3. During the baseline survey, all participants were interviewed face-to-face by trained investigators to complete a questionnaire regarding their socio-demographic characteristics (such as sex, age, ethnicity, family pedigree, marital status, education, occupation, and family income), medical history, lifestyle factors (such as physical activity, dietary behaviours). Smoking history was defined as the participants took at least 100 cigarettes in the past, and drinking was defined as they drank more than 12 times in the past 12 months. Most of these factors are associated with cardiovascular health according to our previous findings [28].

**Table 3.**
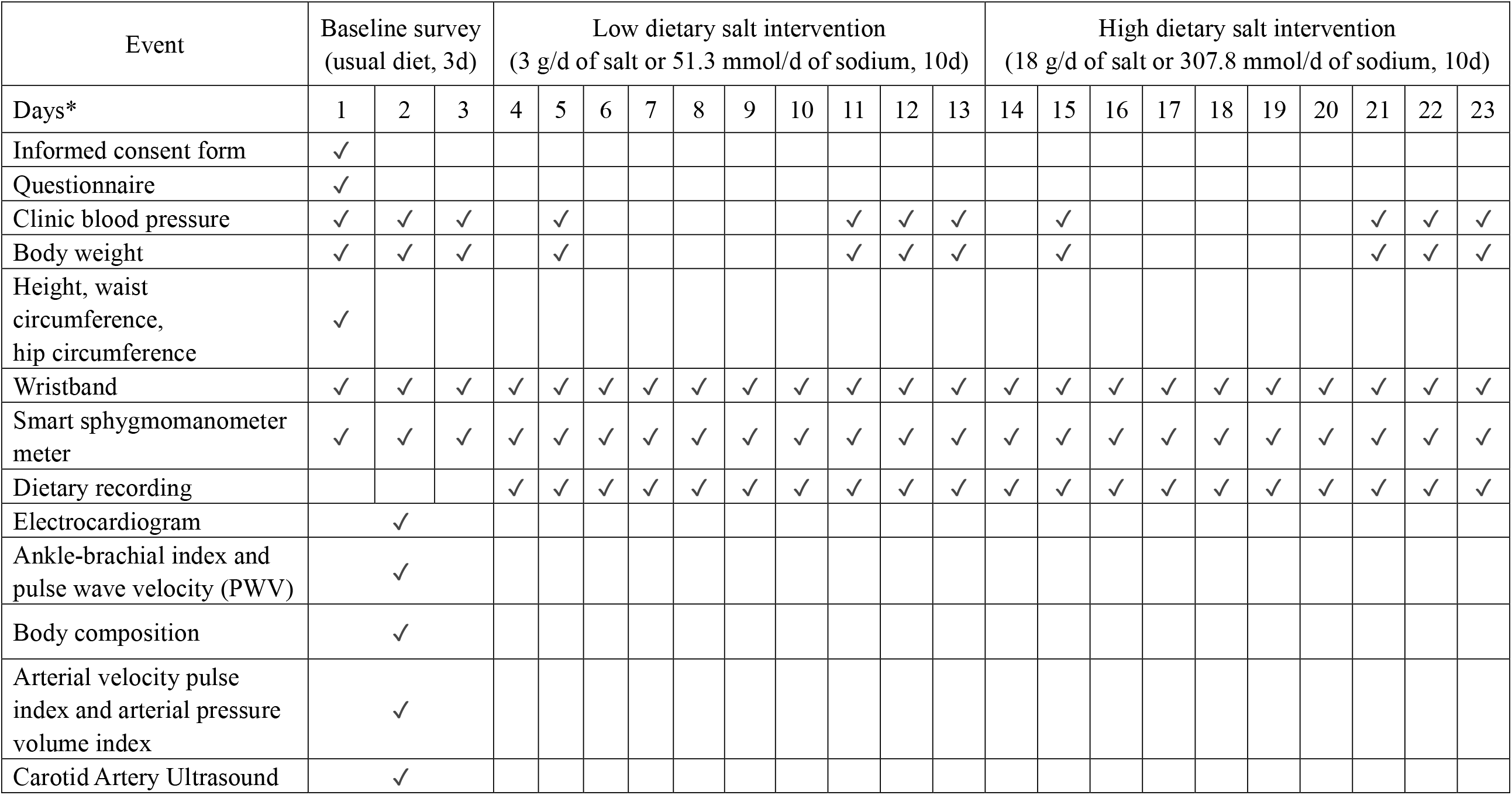
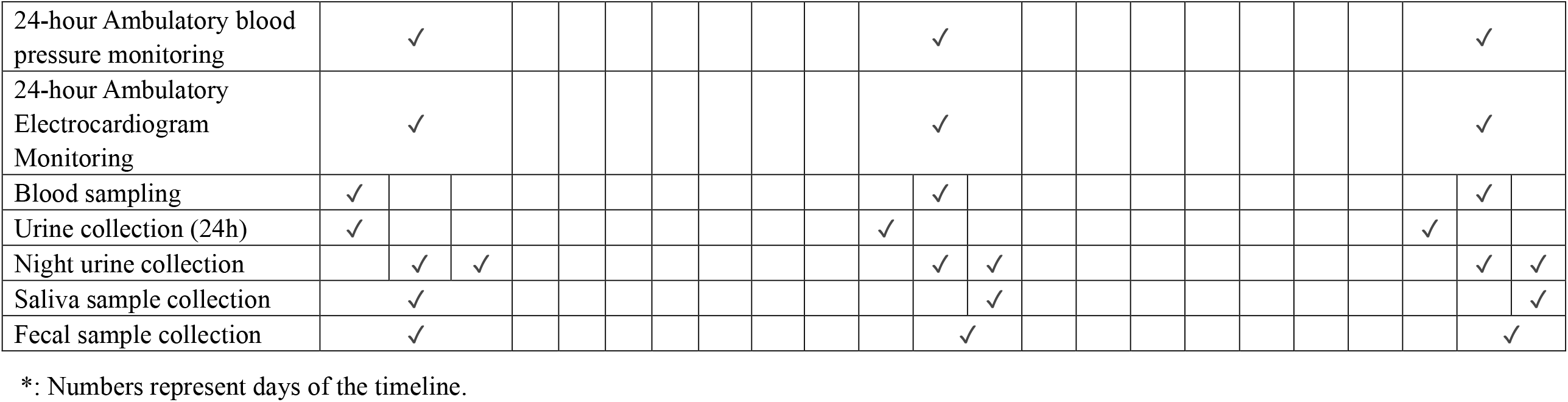
Schedule of the MetaSalt study.

BP was measured three times at each clinical visit by trained and certified staff, in accordance with the instructions of the professional BP monitor (Omron® HBP-1300). Before the measurements, the participants were instructed to stay away from smoking, exercising, eating, taking medicine or drinking caffeine-containing beverages such as tea, coffee and colas for at least 60 minutes prior to the measurement, and keep in the sitting position in a warm and quiet room for five minutes of rest. Moreover, for each day during the study period, BP was also measured at least one time in the morning (after morning urine and before breakfast), one time before lunch, and one time in the night (before sleep) by themselves through a smart sphygmomanometer meter (Lifesense® i5), which could automatically send the measurement data to the staff. Furthermore, 24-hour ambulatory blood pressure monitoring (ABPM) was conducted using KANG® KC-2300A, which was certified by the standards of the Association for the Advancement of Medical Instrumentation (AAMI) or the British Hypertension Society (BHS).

In addition, various parameters were measured using simple non-invasive devices (Table 3): Resting electrocardiogram (ECG) was determined using FUKUDA® 12-Lead Electrocardiography, while a 12-Lead Holter ECG Recorder [Jinco® MIC-12H-3S or Biomedical Instruments (BI®) TeleAECG BI9900] was used to monitor the 24-hour ECG; ankle-brachial index (ABI) and pulse wave velocity (PWV) were determined using Omron® BP-203RPEIII; PASESA® Portable Cardiovascular Measuring Instrument was used to measure arterial velocity pulse index (AVI) and arterial pressure volume index (API); body composition and body fat percentage were determined using Tanita® TBF-418B Professional Body Analyser; Carotid Artery Ultrasound tests were performed using Philips® CX30 Portable Ultrasound System.

We also conducted other anthropometry measurements such as body weight, height, waist and hip circumference, etc. The participants were required to wear a smart wristband (Lifesense mambo HR2) to collect data on their sport, sleep and heart rate during the entire study period.

### Sample collection and preparation

Fasting whole blood was sampled in both Ethylene Diamine Tetraacetic Acid (EDTA) and non-additive blood collection tubes, the serum and plasma were separately stored at −80°C until assayed. Moreover, at the baseline period and each intervention phase, one 24h and two overnight (8h) urine samples were collected for each participant and stored at −20°C until assayed. In addition, at each stage, we used Genotek OMR-501 Saliva Microbiome DNA Collection Kit to collected the saliva samples and Genotek OMR-200 Gut Microbiome DNA Collection Kit to collect the fecal samples, then stored at −80°C until assayed. Untargeted metabolomics profiling will be analysed using serum and urine samples, and the compositions of oral and gut microbiota will be determined with saliva and fecal samples, respectively.

### Quality control

For successful implementation of the MetaSalt study, a series of quality control measures were taken. All study staff was trained by the training program which instructed them to address matters needing attention in conducting and interpreting the face-to-face interviews. The study staff also had to be well acquainted with the questionnaires, measurements, sample collection, methodologies and the operation of the instruments, and use unified criteria for each participant during the investigation. BP was measured using the standard electronic sphygmomanometer, and all observers, who were blinded to the intervention, must attend the special training and pass the examination on knowledge of standard techniques for BP measurement prior to the study. After each interview, all data of the questionnaires were inputted to the MetaSalt study database and subjected to a check for completeness, followed by another round of review at the coordinating centre in Beijing. The discrepancies and illogicality data would be sent back to the study sites to make a correction according to original records or via additional connections. In order to improve the participants’ compliance, the study staff organized and held meetings with the locals to introduce the aims and process of the MetaSalt study, as well as carry out adequate propaganda through leaflets, handbooks and local broadcasts before the baseline survey. During the entire study period, all participants were required to have breakfast, lunch and dinner at specified canteens under the supervision of the investigators, and their food consumption at each meal was recorded. Additionally, an independent coordination centre was set up to monitor and review the entire field investigation, and advise the Steering Committee to modify the trial should it become necessary.

### Data analysis

The characteristics of participants are described as N (%) for categorical variables, and mean (SD) for continuous variables. Percentage changes of the mean levels of mean arterial pressure (MAP), systolic BP (SBP) and diastolic BP (DBP) between baseline and the low-salt intervention period, and between the low- and high-salt intervention periods were calculated, and the differences were compared by paired Student’s *t*-test. The statistical analyses were conducted using R version 3.6.3, and a two-tailed *p*-value below 0.05 will be considered statistically significant.

## Results

The MetaSalt study was conducted from August to November in 2019. A total of 529 subjects from four sites were recruited, included 359 probands and 170 their family members. Among these participants, 512 finished the low salt intervention, and 503 went through the entire study phases with a questionnaire and anthropometric data, and samples (blood, saliva, excrement and urine).

Table 4 shows the baseline characteristics of the study subjects. The mean (SD) of age of all participants was 48.1 (9.3) years old, with 47.3 (10.0) and 48.5 (8.8) years old for the probands and their family members, respectively. We found that 36.7% of the participants were male, 76.5% had a middle school (69.5%) or higher (7.0%) diploma, 23.4% had a history of smoking, 24.4% were current drinkers. They had a BMI of 26.5 (3.6) kg/m^2^ and a waist circumference of 88.6 (9.6) cm. In addition, the mean (SD) heart rate of the entire study population was 75.2 (9.9) beats per minute.

**Table 4.** Baseline characteristics of the participants.

|  | All<br>(n=529) | Probands<br>(n=359) | Family<br>members<br>(n=170) |
| --- | --- | --- | --- |
| Age (SD), years | 48.1 (9.3) | 47.3 (10.0) | 48.5 (8.8) |
| Male (%) | 194 (36.7) | 136 (37.9) | 58 (34.1) |
| Education (%) |  |  |  |
| Primary or lower | 123 (23.4) | 78 (21.9) | 45 (26.6) |
| Middle school | 365 (69.5) | 248 (69.7) | 117 (69.2) |
| Higher | 37 (7.0) | 30 (8.4) | 7 (4.1) |
| History of smoking (%) | 123 (23.4) | 86 (24.1) | 37 (21.9) |
| Drinking (%) | 128 (24.4) | 259 (72.5) | 138 (82.1) |
| Body mass index (SD) , kg/m<sup>2</sup> | 26.5 (3.6) | 27.0 (3.6) | 25.5 (3.4) |
| Waist circumference (SD) , cm | 88.6 (9.6) | 90.0 (9.8) | 85.7 (8.6) |
| Heart rate (SD), beats per minute | 75.2 (9.9) | 75.5 (9.5) | 74.7 (10.7) |

The mean SBP, DBP and MAP levels of the participants at baseline, and during the low-salt and high-salt interventions are showed in Table 5. All of these measurements were decreased during the low-salt intervention in comparing with baseline and elevated during the high-salt intervention in comparing with low-salt intervention. For example, the mean SBP, DBP and MAP levels were 129.54±13.66, 81.02±9.82, and 97.19±10.51 at baseline; 124.50±12.00, 78.41±8.81, and 93.77±9.35 during the low-salt intervention; 129.18±13.61, 80.06±9.16, and 96.43±10.03 during the high-salt intervention, respectively. In comparing with the baseline period, the mean SBP, DBP and MAP of all participants during the low-salt intervention were decreased by 3.89%, 3.22% and 3.52%, respectively. The corresponding increases of 3.76%, 2.10% and 2.84% were observed from the low-salt intervention to the high-salt intervention.

**Table 5.**
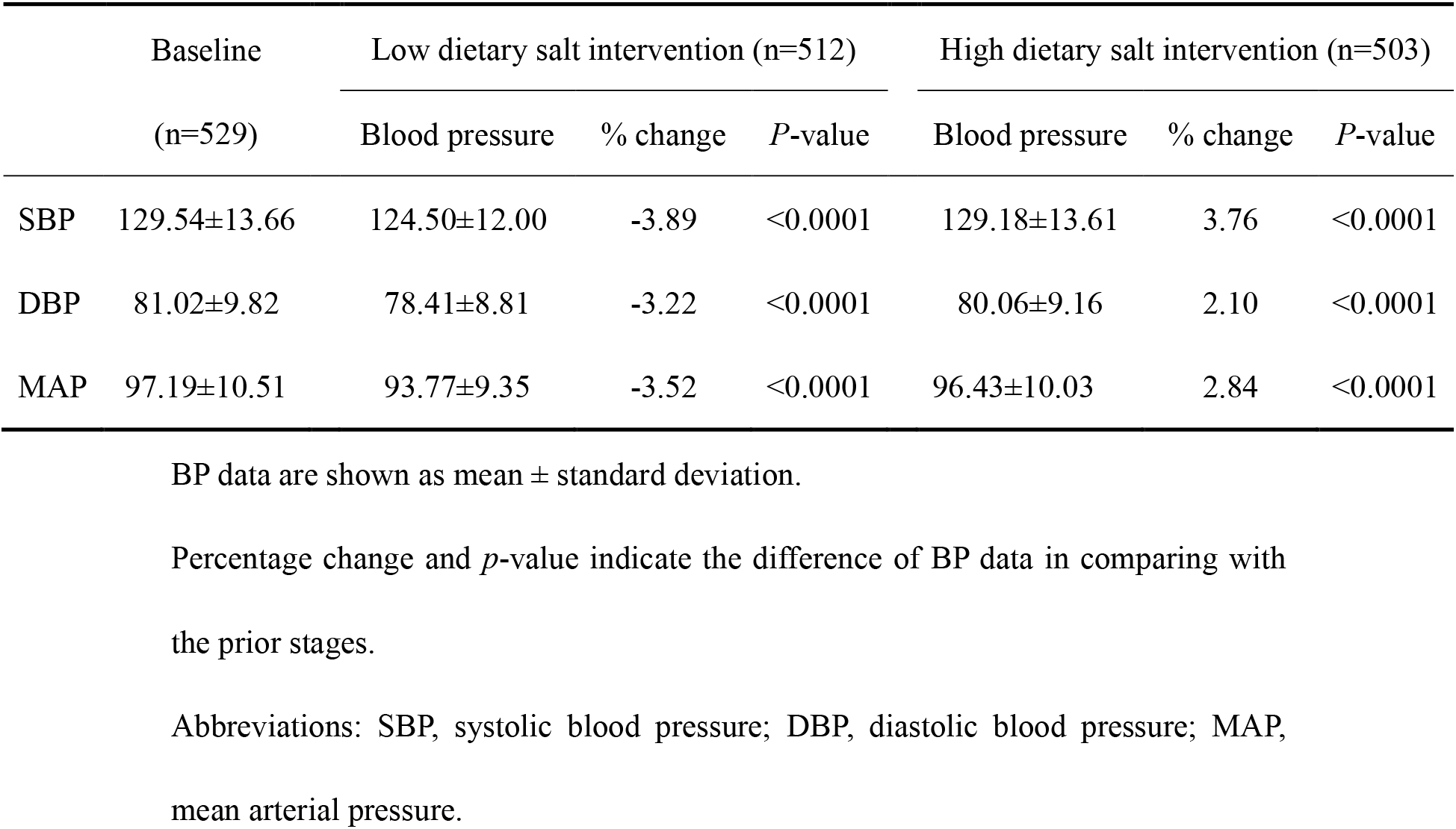
Mean systolic and diastolic BP levels, and mean arterial pressure of the participants during the study.

|  | Baseline<br>(n=529) | Low dietary salt intervention (n=512) |  |  | High dietary salt intervention (n=503) |  |  |
| --- | --- | --- | --- | --- | --- | --- | --- |
|  |  | Blood pressure | % change | *P*-value | Blood pressure | % change | *P*-value |
| SBP | $129.54 \pm 13.66$ | $124.50 \pm 12.00$ | -3.89 | <0.0001 | $129.18 \pm 13.61$ | 3.76 | <0.0001 |
| DBP | $81.02 \pm 9.82$ | $78.41 \pm 8.81$ | -3.22 | <0.0001 | $80.06 \pm 9.16$ | 2.10 | <0.0001 |
| MAP | $97.19 \pm 10.51$ | $93.77 \pm 9.35$ | -3.52 | <0.0001 | $96.43 \pm 10.03$ | 2.84 | <0.0001 |

BP data are shown as mean ± standard deviation.
Percentage change and *p*-value indicate the difference of BP data in comparing with the prior stages.
Abbreviations: SBP, systolic blood pressure; DBP, diastolic blood pressure; MAP, mean arterial pressure.

## Discussion

The MetaSalt study has successfully recruited 529 participants and conducted both low- and high-salt intervention. The SBP, DBP and MAP levels of the participants were significantly decreased in the low-salt intervention, and increased in the high-salt intervention. We also conducted a range of measurements concerning the research questions, and recorded all relevant datasets during every stage of the study, which could enable us to investigate the effects of dietary sodium intake on microbial and metabolomic profiles, as well as subsequent cardiovascular outcomes.

Previous findings have indicated that gut microbiota and metabolomic profiles might play important roles in the association between dietary sodium intake and BP, which might further translate into substantial impacts on cardiovascular health [29, 30]. For example, recent evidence indicated that high salt diets could damage intestinal anatomy and promote tissue inflammation, and the gut microbiota are possible mediators of these inflammatory responses that contributes to the pathogenesis of hypertension [31]. Moreover, there is strong evidence that changes in the circulating metabolome and intestinal microbiota were closely associated with cardiovascular health status, and microbial and metabolomic profiling can be used to identify the crucial mechanisms responsible for the long-term outcomes [17, 18, 32]. Previous findings also suggested that high sodium intake could promote BP elevation by reducing the arachidonic acid and B fragilis levels in the intestine that could further increase the intestinal-derived corticosterone level in the serum and intestine [33]. However, previous Chinese epidemiologic studies were based on an observational design, which were not able to determine the causal relationship. Therefore, the MetaSalt study adopted an intervention design, and collected the blood, urine, saliva and fecal samples, which is of great scientific importance for assessing the mechanisms of the effects of low- and high-dietary sodium intake on BP by focusing on the changes of microbial and metabolomic profiles.

Based on the GenSalt study, which focused on identifying genes related to BP responses to dietary sodium and potassium intake [7], the MetaSalt study was aimed to study the impacts of low- and high-dietary sodium intake on the microbial and metabolomic profiling and subsequent cardiovascular outcomes. These two studies adopted a similar protocol of using a family-based intervention design, but the MetaSalt study had a longer intervention duration of ten days of each phase, whereas the GenSalt study only had seven days [7, 34]. In addition, the mean age of participants in the MetaSalt study was older than those who went through the low- and high-salt intervention in the GenSalt study, and we observed similar patterns of BP decrease during the low-salt intervention and increase during the high-salt intervention [34].

The MetaSalt study has several strengths. First, the intervention design is a distinct advantage for establishing the causal association between the intervening measures and the outcome of interest. Second, the dietary sodium intake of each meal was strictly controlled by pre-packaged salt, and all participants were required to take each of their meals at fixed canteens during the entire study period, with the food consumption recorded, which enable us to measure the effects of dietary sodium intake more accurately. Another merit of the MetaSalt study included the use of several smart devices, such as the smart sphygmomanometer meter and the wearable smart wristband. In addition, although a few participants quitted our study during the interventions, we still kept a high retention rate due to the family-based recruitment of the participants.

In conclusion, the MetaSalt study was well conducted to address an important public health question that how the dietary sodium intake affects microbial and metabolomic profiles. Findings from this study will greatly advance the scientific evidence base for the mechanisms of sodium reduction on cardiovascular health, and therefore may help prioritize strategies for the prevention of cardiovascular diseases.

## Data Availability

The raw data cannot be shared at this time as the data also forms part of an ongoing study.

## Acknowledgments

This work was supported by the Chinese Academy of Medical Sciences (CAMS) Innovation Fund for Medical Sciences (2019-I2M-2-003, 2017-I2M-1-004).

